# Automated Detection and Quantification of Hemorrhagic Transformation After Endovascular Thrombectomy

**DOI:** 10.64898/2026.03.16.26347868

**Authors:** Wi-Sun Ryu, Hee-Jung Ha, Leonard Sunwoo, Myung Jae Lee, Kyusik Kang, Jae Guk Kim, Soo Joo Lee, Jae-Kwan Cha, Tai Hwan Park, Jeong-Yoon Lee, Kyungbok Lee, Doo Hyuk Kwon, Jun Lee, Hong-Kyun Park, Keun-Sik Hong, Minwoo Lee, Mi Sun Oh, Kyung-Ho Yu, Dong-Seok Gwak, Dong-Eog Kim, Hyunsoo Kim, Joon-Tae Kim, Joong-Goo Kim, Jay Chol Choi, Wook-Joo Kim, Jee Hyun Kwon, Kyu Sun Yum, Dong-Ick Shin, Jeong-Ho Hong, Sung-Il Sohn, Sang-Hwa Lee, Chulho Kim, Hae-Bong Jeong, Kwang-Yeol Park, Keon-Joo Lee, Chi Kyung Kim, Jihoon Kang, Jun Yup Kim, Hee-Joon Bae, Beom Joon Kim

## Abstract

**Background:** Hemorrhagic transformation (HT) after endovascular thrombectomy (EVT) is a principal determinant of clinical outcome. Artificial intelligence (AI) algorithms for spontaneous hemorrhage detection exist, but none has been validated for post-procedural HT across multiple imaging modalities.

**Methods:** We conducted a multicenter diagnostic accuracy study within the Clinical Research Collaboration for Stroke in Korea registry (18 centers, 2022–2023). Patients who underwent EVT and received follow-up NCCT, GRE, or SWI within 168 hours were included. AI-derived hemorrhage volumes were compared against expert-determined ECASS classification. Three-month modified Rankin Scale (mRS) scores were evaluated for volume–outcome association.

**Results:** Among 1,490 patients (median age 73; 57.4% male), HT was present in 41.4% and parenchymal hemorrhage (PH) in 11.1%. PH detection sensitivity exceeded 94% across all modalities (NCCT 95.4%, GRE 94.4%, SWI 98.3%), with AUCs of 0.900, 0.943, and 0.953, respectively. AI-derived volume correlated with 3-month mRS (Spearman ρ = 0.353, P < 0.001); good outcome (mRS 0–2) declined from 61.8% to 6.7% across increasing volume categories. Among ECASS 0 cases, AI-positive patients had significantly worse outcomes than true-negatives (good outcome 48.2% vs 67.2%, mortality 10.7% vs 4.6%, P < 0.001).

**Conclusions:** AI-based hemorrhage quantification provides high detection of clinically significant PH after EVT and demonstrates a dose–response association with functional outcome. AI-derived volume may serve as a continuous prognostic biomarker that identifies at-risk subgroups beyond categorical ECASS grading.

## INTRODUCTION

The clinical management of acute ischemic stroke has been transformed by the advent of endovascular thrombectomy (EVT) [1,2]. The restoration of blood flow to ischemic tissue inherently predisposes it to hemorrhagic transformation (HT), a complication spanning from benign petechial staining to space-occupying hematoma [3,4]. While the manual grading of HT by European Cooperative Acute Stroke Study (ECASS) or Heidelberg Bleeding Classification (HBC) is accepted reference standard for classification, [5,6] the expansion of multicenter registries to thousands of studies renders centralized expert adjudication impractical at scale. [7,8]

Although AI algorithms for spontaneous hemorrhage detection have achieved high diagnostic accuracy [9,10], the post-procedural setting presents distinct radiographic confounders. Iodinated contrast staining, ischemic edema, and blood-brain barrier disruption create mixed-density appearances that may not be represented in the training data of most commercial algorithms. [11,12]. Furthermore, the application of AI to magnetic resonance imaging (MRI) sequences, specifically gradient-recalled echo (GRE) and susceptibility-weighted imaging (SWI), remains unexplored. Given the superior sensitivity of MRI to blood products, MR tends to be the prioritized follow-up modality in several specialized stroke centers [13,14].

In this multicenter study, we analyzed a nationwide prospective database from 18 university hospitals [7,15] to evaluate the diagnostic fidelity of AI-based hemorrhage quantification across NCCT, GRE, and SWI. Utilizing expert ECASS classification as our reference standard, we delineate binary detection performance and the AI-derived volumetric thresholds that optimize the identification of clinically significant parenchymal hemorrhage (PH), and to examine whether algorithmically detected signals among expert-classified no-HT cases carry independent prognostic significance.

## METHODS

### Data Availability and Reporting Guidelines

Data supporting the findings of this study are available from the corresponding author upon reasonable request from qualified researchers, subject to privacy and ethical restrictions. This study was conducted in accordance with the Standards for Reporting of Diagnostic Accuracy Studies (STARD).

### Ethics Approval and Informed Consent

The Clinical Research Collaboration for Stroke in Korea (CRCS-K) registry and the present analysis were approved by the institutional review boards of all participating centers. Written informed consent for registry enrollment and for the use of clinical and imaging data for research purposes was obtained from all patients or their legally authorized representatives. For the current study, which used only deidentified data extracted from the CRCS-K registry, the protocol was additionally reviewed and approved by the institutional review board of Seoul National University Bundang Hospital, the coordinating center (IRB No. B-2308-845-302). All procedures were conducted in accordance with the Declaration of Helsinki.

### Study design and population

The present investigation was performed through a retrospective analysis of clinical and neuroimaging data obtained from the CRCS-K Image Repository. This nationwide and multicenter clinical and imaging registry was established to systematically collect and archive all stroke images obtained during patient admissions since its inception in June 2022 across eighteen participating academic institutions. The protocol was approved by institutional review boards of all participating centers and conducted in accordance with the Standards for Reporting of Diagnostic Accuracy Studies guidelines [16].

Patients were eligible for inclusion if they underwent EVT for anterior circulation large-vessel occlusion between June 2022 and December 2023 and received at least one follow-up brain imaging study (NCCT, GRE, or SWI) within 168 hours (7 days) of the index procedure. This time window was selected to encompass the clinically relevant period during which the majority of HT develop and are detected [3]. When multiple imaging modalities were available for a single patient, the more sensitive magnetic resonance sequences including GRE or SWI were prioritized over NCCT. Patients were excluded if imaging was of insufficient quality for interpretation, if no DICOM data were available for AI processing, or if the HT was identified beyond the 168-hour window.

### Reference standard: ECASS hemorrhagic transformation grading

Follow-up imaging was performed according to individual center practices using SWI or GRE or NCCT. Hemorrhagic transformation was defined as the presence of susceptibility hypointensity signals on magnetic resonance imaging or hyperdensity lesions on NCCT followed as ECASS criteria [5,6]. For the purposes of diagnostic performance analysis, two clinically relevant binary endpoints were defined: any hemorrhagic transformation (ECASS ≥1 versus ECASS 0) and PH (ECASS ≥3 versus ECASS 0–2). All scans were independently reviewed at a centralized core image laboratory by two vascular experts who remained blinded to all clinical characteristics and functional outcomes. The inter-rater agreement was a κ of 0.867 for binary detection of any hemorrhage. For the classification of patients into categories of no hemorrhage versus hemorrhagic infarction versus PH the quadratically weighted κ was 0.933 while the agreement for the full strata of transformation reached 0.894. Any disagreements between the primary reviewers were resolved through a consensus discussion.

### AI Hemorrhage Quantification Algorithms

#### NCCT algorithm

For NCCT, hemorrhage detection and volumetric quantification were performed using a commercially available, regulatory-cleared deep learning algorithm (JLK Inc., Seoul, South Korea) previously validated for spontaneous intracranial hemorrhage with a sensitivity of 98.7% and area under the curve of 0.936 in an independent multicenter evaluation [9]. The algorithm was applied without retraining or domain adaptation; the present study constitutes its first systematic evaluation in the post-EVT hemorrhagic transformation setting.

#### GRE and SWI algorithms

For GRE (T2*-weighted) and SWI sequences, dedicated segmentation models were developed using the nnU-Net framework, a self-configuring method for deep learning–based biomedical image segmentation [17]. Both models employ a 3D fully convolutional encoder–decoder architecture with skip connections and perform voxel-level multi-label segmentation of three classes: hemorrhage, superficial siderosis, and cerebral microbleeds. The preprocessing pipeline comprised SynthStrip-based skull stripping [18], resampling to dataset-derived target spacing, z-score intensity normalization, and extraction of overlapping 3D patches (Supplementary Methods; Supplementary Table 1). Models were trained with a combined Dice and binary cross-entropy loss using the Adam optimizer (learning rate 3×10⁻□). The GRE model was trained on 1,441 multi-vendor studies from 12 institutions and externally validated on 310 independent studies from 2 institutions not represented in training, achieving a Dice similarity coefficient of 0.938. The SWI model was trained on 1,152 studies from 12 institutions and validated on 216 independent studies (DSC 0.862). All training annotations were generated at the voxel level and adjudicated by two board-certified vascular neurologists. Full details of the network architecture, hyperparameters, and imaging parameter distributions are provided in the Supplementary Methods and Supplementary Tables 1–2.

To differentiate PH from superficial siderosis, a separately trained 2D CSF segmentation model was applied post hoc: segmented components with ≥50% spatial overlap with CSF regions were reclassified as siderosis and excluded from the hemorrhage volume (Supplementary Figure 1).

#### AI output definition

All imaging studies were processed in batch mode without manual intervention. The AI output for each study consisted of a binary detection result and a continuous volumetric measurement in milliliters. An AI-positive result was defined as hemorrhage volume exceeding 0 mL.

### Clinical outcome assessment

Functional outcome was assessed using the modified Rankin Scale (mRS) at 3 months, obtained through clinic visits or structured telephone interviews per the CRCS-K follow-up protocol. Good outcome was defined as mRS 0–2, and mortality as mRS 6. To evaluate the prognostic salience of AI-derived volume, patients were stratified into five clinically informed categories (0, 0–1.6, 1.6–10, 10–50, and >50 mL) anchored to the optimal thresholds from the diagnostic accuracy analysis. ECASS 0 cases were further dichotomized into true-negative (AI volume = 0 mL) and AI-positive (AI volume > 0 mL) subgroups to determine whether algorithmically detected signals absent expert-adjudicated hemorrhage carried prognostic significance.

### Statistical analysis

Baseline characteristics were summarized as medians with interquartile ranges (IQR) for continuous variables and as counts with percentages for categorical variables. Comparisons across imaging modalities were performed using the Kruskal-Wallis test for continuous variables and the chi-square test for categorical variables.

Diagnostic performance was evaluated at two levels. First, binary detection performance (AI-positive versus AI-negative) was assessed against the ECASS reference standard for both any hemorrhagic transformation and PH endpoints. Sensitivity, specificity, positive predictive value, negative predictive value, and accuracy were calculated with 95% Wilson score confidence intervals. Second, receiver operating characteristic (ROC) analysis was performed using AI-derived hemorrhage volume as a continuous discriminator. The area under the ROC curve (AUC) was estimated with DeLong 95% confidence intervals. Optimal volume thresholds were determined by maximizing the Youden index (sensitivity + specificity − 1). AI detection rates were additionally reported by individual ECASS grade to characterize performance across the spectrum of hemorrhagic transformation severity. All statistical analyses were performed using Python 3.11 (Python Software Foundation) with the scikit-learn, SciPy, and NumPy libraries. The association between AI-derived volume and 3-month mRS was quantified using Spearman rank correlation. Differences in mRS distribution between subgroups were evaluated using the Mann-Whitney U test. A two-sided P value below 0.05 was considered statistically significant.

## RESULTS

### Baseline characteristics of study population

Among 1,565 patients receiving recanalization therapy a total of 1,490 cases were included for final analysis (Figure 1). These included 320 NCCT examinations (21.5%), 420 GRE studies (28.2%), and 750 SWI studies (50.3%). The median age was 73 years (IQR 62–81; Table 1), 855 patients (57.4%) were male, and the median National Institutes of Health Stroke Scale (NIHSS) score at admission was 10 (IQR 5–15). The median Alberta Stroke Program Early CT Score (ASPECTS) was 8 (IQR 7–9). Any HT was observed in 41.4% of patients (n=617) including HI1 in 13.2% (n=196), HI2 in 17.2% (n=256), PH1 in 7.3% (n=109), and PH2 in 3.8% (n=56). Patients receiving NCCT as the follow up modality presented with significantly higher baseline NIHSS scores (median 14.0 versus 9.0 for GRE and 9.0 for SWI, P < 0.001; Supplemental Table 3) and lower initial ASPECTS (median 7 versus 9 for GRE and 9 for SWI, P < 0.001) than those evaluated with MRI based imaging (Supplementary Figure 2).

**Figure 1.**
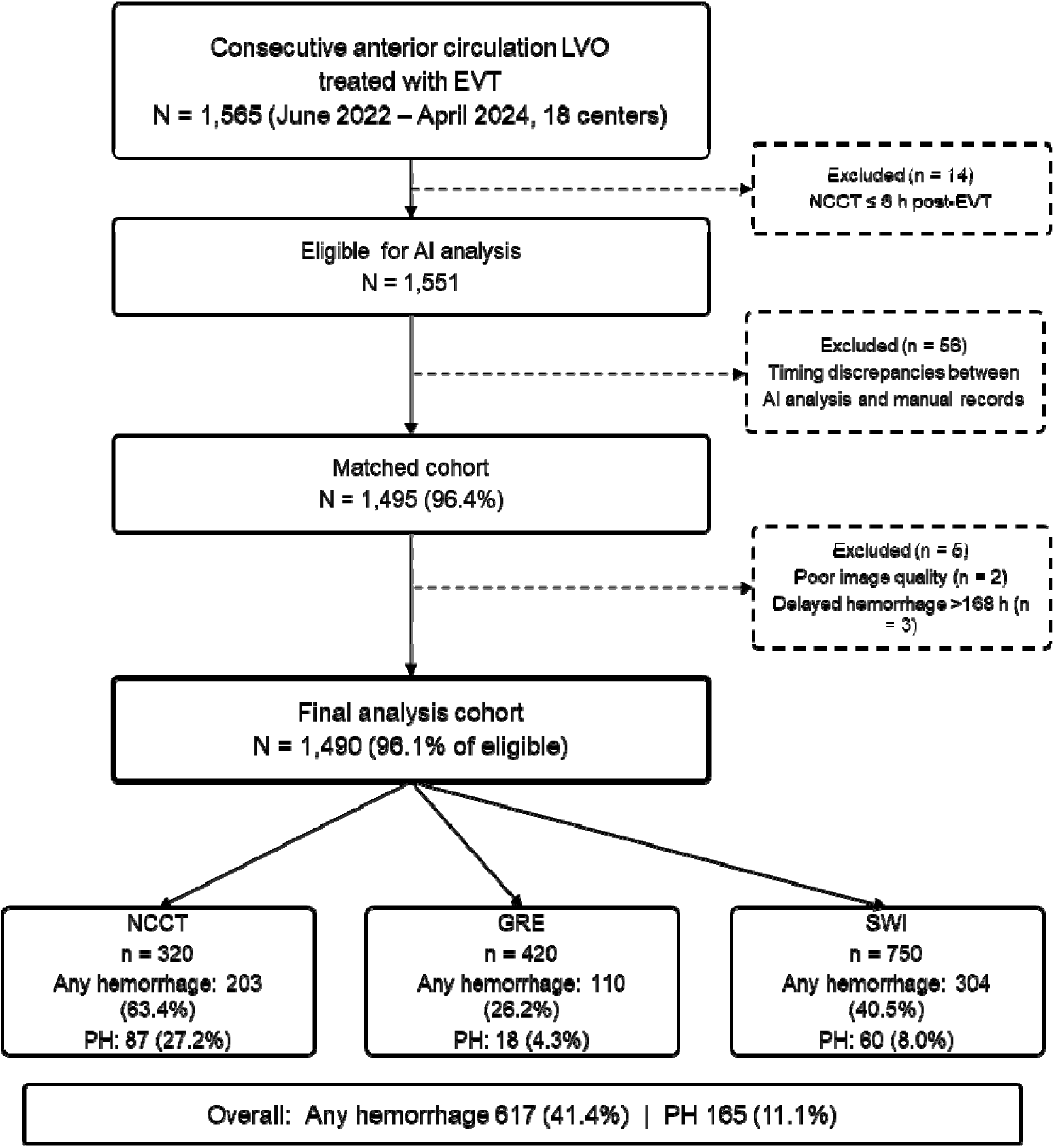
Patient selection flowchart. Abbreviations: NCCT, non-contrast computed tomography; GRE, gradient-recalled echo; SWI, susceptibility-weighted imaging; EVT, endovascular thrombectomy; HT, hemorrhagic transformation; PH, parenchymal hemorrhage.

**Table 1.** Baseline Characteristics of the Study Population by Imaging Modality.

|  | Overall<br>(N=1,490) | NCCT<br>(n=320) | GRE<br>(n=420) | SWI<br>(n=750) | P value |
| --- | --- | --- | --- | --- | --- |
| N | 1490 | 320 | 420 | 750 |  |
| Age, years | 73 (62–81) | 75 (64–82) | 71 (61–80) | 72 (62–80) | 0.004 |
| Male sex, n (%) | 855 (57.4) | 171 (53.4) | 256 (61.0) | 428 (57.1) | 0.137 |
| NIHSS at admission | 10 (5–15) | 14 (8–18) | 9 (5–15) | 9 (5–14) | <0.001 |
| ASPECTS | 8 (7–9) | 7 (5–9) | 9 (7–10) | 9 (7–10) | <0.001 |
| Pre-stroke mRS | 0 (0–0) | 0 (0–1) | 0 (0–0) | 0 (0–0) | <0.001 |
| Comorbidities |  |  |  |  |  |
| Hypertension, n (%) | 917 (61.5) | 206 (64.4) | 277 (66.0) | 434 (57.9) | 0.021 |
| Diabetes mellitus, n (%) | 420 (28.2) | 96 (30.0) | 124 (29.5) | 200 (26.7) | 0.363 |
| Atrial fibrillation, n (%) | 573 (38.5) | 163 (50.9) | 150 (35.7) | 260 (34.7) | <0.001 |
| Dyslipidemia, n (%) | 534 (35.8) | 131 (40.9) | 193 (46.0) | 210 (28.0) | <0.001 |
| Prior stroke, n (%) | 242 (16.2) | 68 (21.2) | 59 (14.0) | 115 (15.3) | 0.035 |
| Current smoker, n (%) | 415 (27.9) | 78 (24.4) | 122 (29.0) | 215 (28.7) | 0.282 |
| IV thrombolysis, n (%) | 986 (66.2) | 178 (55.6) | 294 (70.0) | 514 (68.5) | <0.001 |
| TOAST classification |  |  |  |  |  |
| Large artery atherosclerosis, n (%) | 392 (26.3) | 79 (24.7) | 127 (30.2) | 186 (24.8) |  |
| Cardioembolism, n (%) | 72 (4.8) | 4 (1.2) | 30 (7.1) | 38 (5.1) |  |
| Small vessel occlusion, n (%) | 605 (40.6) | 164 (51.2) | 160 (38.1) | 281 (37.5) |  |
| Other determined, n (%) | 66 (4.4) | 17 (5.3) | 12 (2.9) | 37 (4.9) |  |
| Two or more causes, n (%) | 49 (3.3) | 8 (2.5) | 16 (3.8) | 25 (3.3) |  |
| Undetermined (incomplete), n (%) | 157 (10.5) | 24 (7.5) | 47 (11.2) | 86 (11.5) |  |
| Undetermined (unclassified), n (%) | 75 (5.0) | 19 (5.9) | 4 (1.0) | 52 (6.9) |  |
| Undetermined (negative), n (%) | 73 (4.9) | 5 (1.6) | 23 (5.5) | 45 (6.0) |  |
| Hemorrhagic outcomes |  |  |  |  |  |
| Any hemorrhage (ECASS $\geq 1$), n (%) | 617 (41.4) | 203 (63.4) | 110 (26.2) | 304 (40.5) | |
| PH (ECASS $\geq 3$), n (%) | 165 (11.1) | 87 (27.2) | 18 (4.3) | 60 (8.0) | |
| Imaging timing, hours after EVT | 37.5<br>(21.5–91.2) | 33.7<br>(21.1–83.8) | 24.0<br>(18.8–77.2) | 56.0<br>(24.0–96.0) | <0.001 |
| 3-month mRS | 2 (1–4) | 4 (2–5) | 2 (1–4) | 2 (1–4) | <0.001 |

Values are median (IQR) or n (%). P values from Kruskal-Wallis or chi-square tests. Abbreviations: EVT, endovascular thrombectomy; NIHSS, National Institutes of Health Stroke Scale; ASPECTS, Alberta Stroke Program Early CT Score; mRS, modified Rankin Scale; TOAST, Trial of Org 10172 in Acute Stroke Treatment; PH, parenchymal hemorrhage; ECASS, European Cooperative Acute Stroke Study; IQR, interquartile range.

### Diagnostic Performance for Hemorrhagic Transformation

Diagnostic performance was evaluated for two pre-specified endpoints; any HT (ECASS ≥1) and parenchymal hemorrhage (ECASS ≥3). In the detection of any HT (Panel A), the sensitivity ranged from 61.8% for GRE to 79.6% for SWI while specificity ranged from 83.8% for NCCT to 90.6% for GRE (Table 2). For the PH detection (Panel B) the sensitivity was 95.4% (95% CI 88.8 to 98.2) for NCCT, 94.4% (95% CI 74.2 to 99.0) for GRE, 98.3% (95% CI 91.1 to 99.7) for SWI. Corresponding specificities were 66.1% (95% CI 59.8 to 71.9), 80.1% (95% CI 75.8 to 83.7), and 64.1% (95% CI 60.4 to 67.6), respectively. Detection rates for the algorithm by grade as NCCT identified 16.2% of cases with no hemorrhage, 39.3% of HI1, 55.7% of HI2, 92.0% of PH1, and 100.0% of PH2. A similar gradient was shown for GRE (9.4%, 25.5%, 92.7%, 100.0%, and 83.3%) and SWI (14.6%, 60.7%, 88.2%, 97.9%, and 100.0%) (Figure 2).

**Figure 2.**
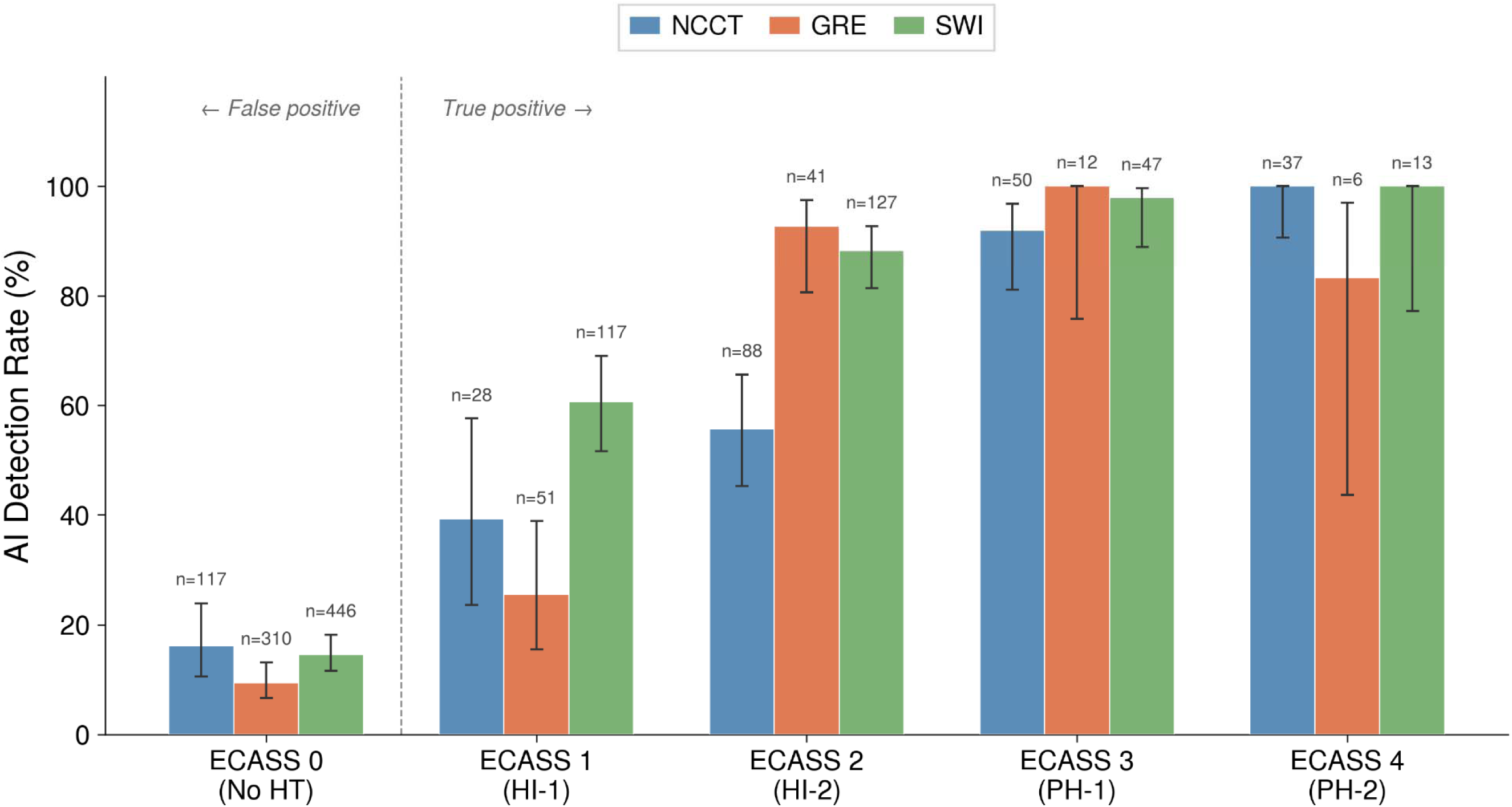
AI-positive detection rates by ECASS hemorrhagic transformation grade and imaging modality. Detection rates increase monotonically with hemorrhage severity across all three modalities, with near-complete detection achieved for PH-2 (ECASS 4). Abbreviations: AI, artificial intelligence; ECASS, European Cooperative Acute Stroke Study; GRE, gradient-recalled echo; HI, hemorrhagic infarction; NCCT, non-contrast computed tomography; PH, parenchymal hemorrhage; SWI, susceptibility-weighted imaging.

**Table 2.**
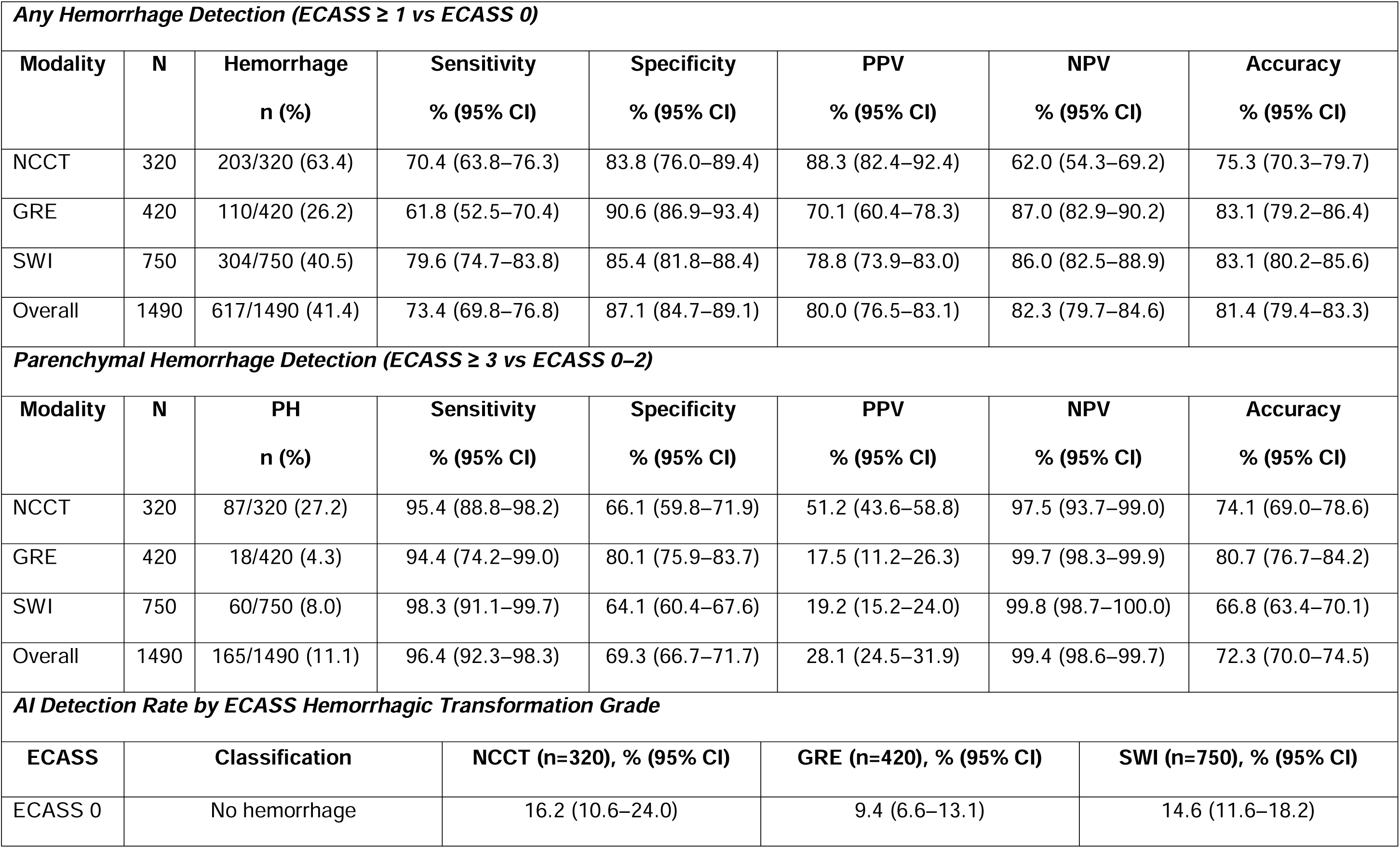
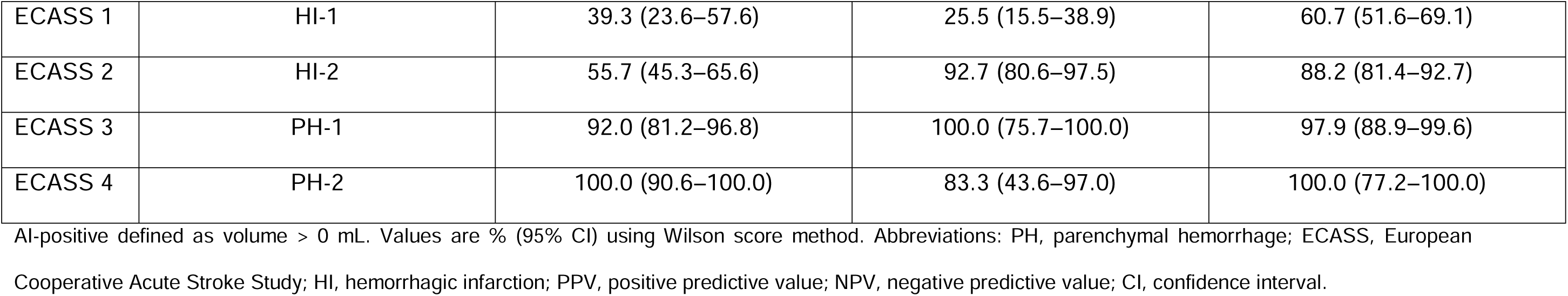
Diagnostic Performance of AI Hemorrhage Quantification by Imaging Modality.

| ***Any Hemorrhage Detection (ECASS $\geq 1$ vs ECASS 0)*** |  |  |  |  |  |  |  |
| --- | --- | --- | --- | --- | --- | --- | --- |
| **Modality** | **N** | **Hemorrhage**<br>**n (%)** | **Sensitivity**<br>**% (95% CI)** | **Specificity**<br>**% (95% CI)** | **PPV**<br>**% (95% CI)** | **NPV**<br>**% (95% CI)** | **Accuracy**<br>**% (95% CI)** |
| NCCT | 320 | 203/320 (63.4) | 70.4 (63.8–76.3) | 83.8 (76.0–89.4) | 88.3 (82.4–92.4) | 62.0 (54.3–69.2) | 75.3 (70.3–79.7) |
| GRE | 420 | 110/420 (26.2) | 61.8 (52.5–70.4) | 90.6 (86.9–93.4) | 70.1 (60.4–78.3) | 87.0 (82.9–90.2) | 83.1 (79.2–86.4) |
| SWI | 750 | 304/750 (40.5) | 79.6 (74.7–83.8) | 85.4 (81.8–88.4) | 78.8 (73.9–83.0) | 86.0 (82.5–88.9) | 83.1 (80.2–85.6) |
| Overall | 1490 | 617/1490 (41.4) | 73.4 (69.8–76.8) | 87.1 (84.7–89.1) | 80.0 (76.5–83.1) | 82.3 (79.7–84.6) | 81.4 (79.4–83.3) |
| ***Parenchymal Hemorrhage Detection (ECASS $\geq 3$ vs ECASS 0–2)*** |  |  |  |  |  |  |  |
| **Modality** | **N** | **PH**<br>**n (%)** | **Sensitivity**<br>**% (95% CI)** | **Specificity**<br>**% (95% CI)** | **PPV**<br>**% (95% CI)** | **NPV**<br>**% (95% CI)** | **Accuracy**<br>**% (95% CI)** |
| NCCT | 320 | 87/320 (27.2) | 95.4 (88.8–98.2) | 66.1 (59.8–71.9) | 51.2 (43.6–58.8) | 97.5 (93.7–99.0) | 74.1 (69.0–78.6) |
| GRE | 420 | 18/420 (4.3) | 94.4 (74.2–99.0) | 80.1 (75.9–83.7) | 17.5 (11.2–26.3) | 99.7 (98.3–99.9) | 80.7 (76.7–84.2) |
| SWI | 750 | 60/750 (8.0) | 98.3 (91.1–99.7) | 64.1 (60.4–67.6) | 19.2 (15.2–24.0) | 99.8 (98.7–100.0) | 66.8 (63.4–70.1) |
| Overall | 1490 | 165/1490 (11.1) | 96.4 (92.3–98.3) | 69.3 (66.7–71.7) | 28.1 (24.5–31.9) | 99.4 (98.6–99.7) | 72.3 (70.0–74.5) |
| ***AI Detection Rate by ECASS Hemorrhagic Transformation Grade*** |  |  |  |  |  |  |  |
| **ECASS** | **Classification** |  | **NCCT (n=320), % (95% CI)** |  | **GRE (n=420), % (95% CI)** |  | **SWI (n=750), % (95% CI)** |
| ECASS 0 | No hemorrhage |  | 16.2 (10.6–24.0) |  | 9.4 (6.6–13.1) |  | 14.6 (11.6–18.2) |

|  |  |  |  |  |
| --- | --- | --- | --- | --- |
| ECASS 1 | HI-1 | 39.3 (23.6–57.6) | 25.5 (15.5–38.9) | 60.7 (51.6–69.1) |
| ECASS 2 | HI-2 | 55.7 (45.3–65.6) | 92.7 (80.6–97.5) | 88.2 (81.4–92.7) |
| ECASS 3 | PH-1 | 92.0 (81.2–96.8) | 100.0 (75.7–100.0) | 97.9 (88.9–99.6) |
| ECASS 4 | PH-2 | 100.0 (90.6–100.0) | 83.3 (43.6–97.0) | 100.0 (77.2–100.0) |

AI-positive defined as volume > 0 mL. Values are % (95% CI) using Wilson score method. Abbreviations: PH, parenchymal hemorrhage; ECASS, European Cooperative Acute Stroke Study; HI, hemorrhagic infarction; PPV, positive predictive value; NPV, negative predictive value; CI, confidence interval.

### Volumetric Analysis and ROC Optimal Thresholds

To identify modality-specific volume thresholds for PH detection, ROC analysis was performed using AI-derived hemorrhage volume as a continuous discriminator. The area under the curve reached 0.900 (95% CI 0.858–0.938) for NCCT, 0.943 (95% CI 0.912–0.974) for GRE, and 0.953 (95% CI 0.933–0.973) for SWI (Figure 3 and Table 3). Optimal volume thresholds for PH detection determined by the Youden index were 1.8 mL for NCCT, 1.6 mL for GRE, and 1.6 mL for SWI. The distribution of detected volumes across individual grades confirmed that median volumes increased progressively with hemorrhage severity (Figure 4).

**Figure 3.**
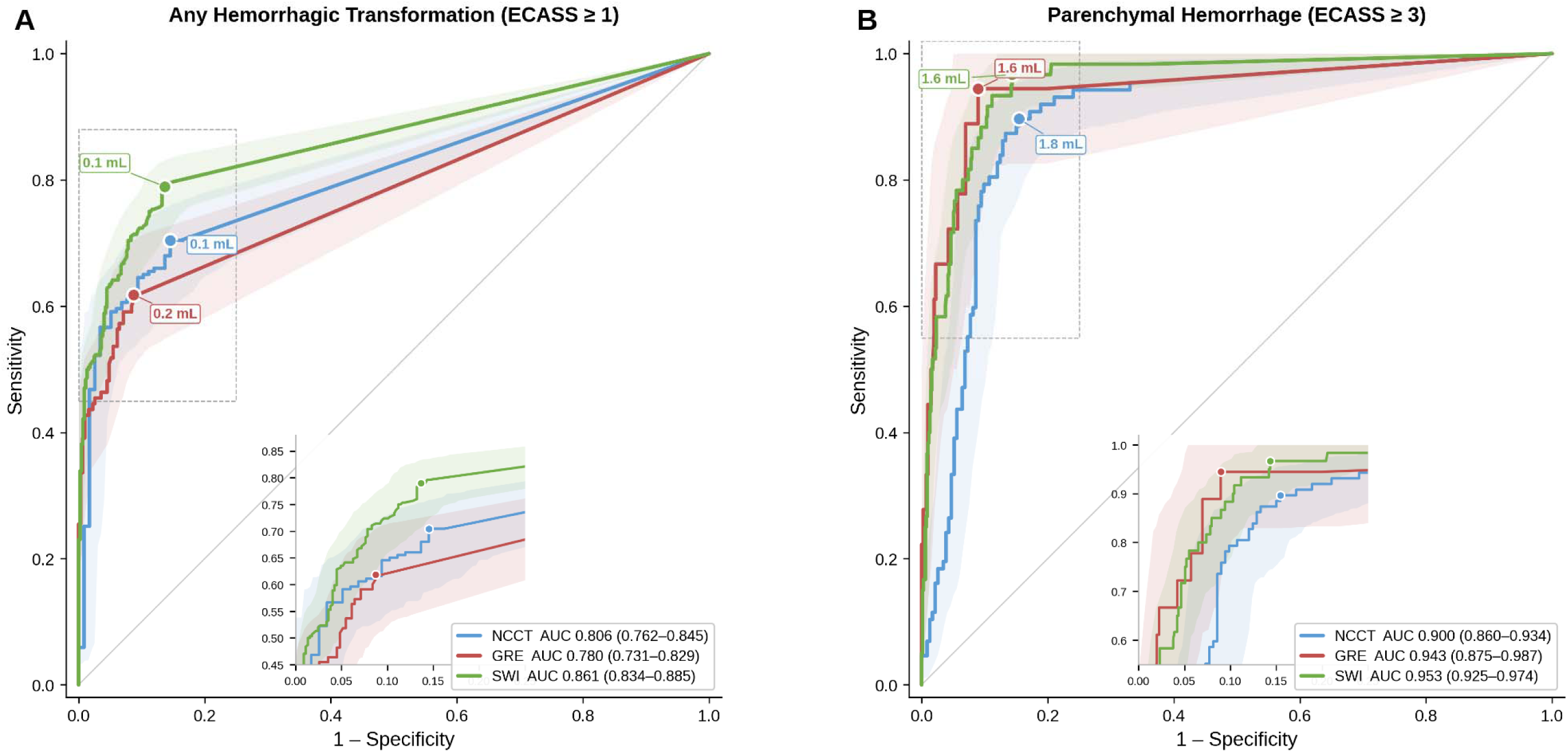
Receiver operating characteristic curves for AI-based hemorrhage detection by imaging modality. Panel A: any hemorrhagic transformation (ECASS ≥1). Panel B: parenchymal hemorrhage (ECASS ≥3). MRI-based modalities (GRE, SWI) achieve superior discrimination compared with NCCT, particularly for the PH endpoint. Abbreviations: AUC, area under the curve; ECASS, European Cooperative Acute Stroke Study; GRE, gradient-recalled echo; NCCT, non-contrast computed tomography; PH, parenchymal hemorrhage; SWI, susceptibility-weighted imaging.

**Figure 4.**
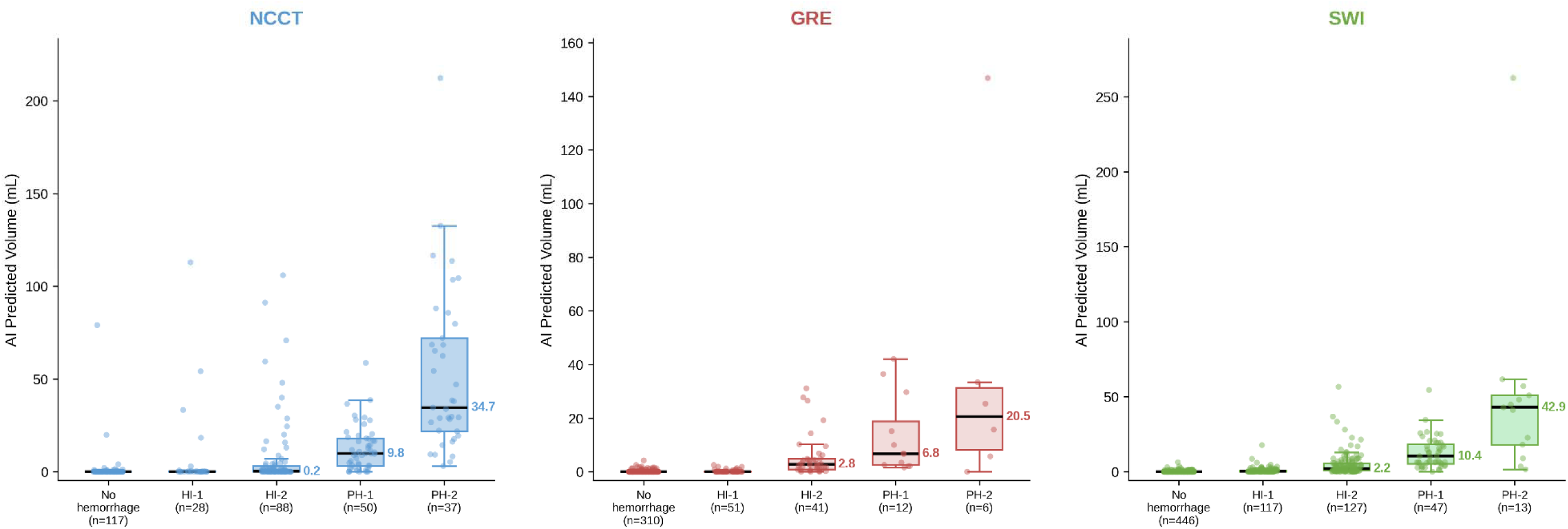
Distribution of AI-derived hemorrhage volumes by ECASS grade and imaging modality. Box plots depict median, interquartile range, and individual outliers. The clear volumetric separation between HI (ECASS 1–2) and PH (ECASS 3–4) categories supports the use of volume-based thresholds for parenchymal hemorrhage detection. Abbreviations: AI, artifici l intelligence; ECASS, European Cooperative Acute Stroke Study; HI, hemorrhagic infarction; IQR, interquartile range; PH, parenchymal hemorrhage.

**Table 3.** AI Volume Thresholds for Parenchymal Hemorrhage Detection by Imaging Modality.

| **NCCT ($n=320$, $PH=87$ [27.2%]) — AUC 0.900 (0.858–0.938)** |  |  |  |  |  |  |  |
| --- | --- | --- | --- | --- | --- | --- | --- |
| **Threshold**<br>**(mL)** | **Sensitivity**<br>**% (95% CI)** | **Specificity**<br>**% (95% CI)** | **PPV**<br>**% (95% CI)** | **NPV**<br>**% (95% CI)** | **Youden**<br>**Index** | **FN**<br>**(missed**<br>**PH)** | **FP**<br>**(ECASS**<br>**0–2)** |
| 0.1 | 94.3 (87.2–97.5) | 67.8 (61.6–73.5) | 52.2 (44.5–59.9) | 96.9 (93.0–98.7) | 0.621 | 5 | 75 |
| 0.5 | 94.3 (87.2–97.5) | 75.5 (69.6–80.6) | 59.0 (50.7–66.8) | 97.2 (93.7–98.8) | 0.698 | 5 | 57 |
| 1.0 | 92.0 (84.3–96.0) | 80.3 (74.7–84.9) | 63.5 (54.8–71.4) | 96.4 (92.7–98.2) | 0.722 | 7 | 46 |
| 1.5 | 90.8 (82.9–95.3) | 82.8 (77.5–87.1) | 66.4 (57.5–74.2) | 96.0 (92.3–98.0) | 0.736 | 8 | 40 |
| **1.8 \*** | **89.7 (81.5–94.5)** | **84.5 (79.4–88.6)** | **68.4 (59.4–76.2)** | **95.6 (91.9–97.7)** | **0.742** | **9** | **36** |
| 2.0 | 88.5 (80.1–93.6) | 85.0 (79.8–89.0) | 68.8 (59.7–76.6) | 95.2 (91.4–97.4) | 0.735 | 10 | 35 |
| 3.0 | 86.2 (77.4–91.9) | 87.1 (82.2–90.8) | 71.4 (62.2–79.2) | 94.4 (90.5–96.8) | 0.733 | 12 | 30 |
| 5.0 | 78.2 (68.4–85.5) | 90.1 (85.6–93.3) | 74.7 (64.9–82.5) | 91.7 (87.4–94.6) | 0.683 | 19 | 23 |
| 10.0 | 64.4 (53.9–73.6) | 91.4 (87.1–94.4) | 73.7 (62.8–82.3) | 87.3 (82.5–90.9) | 0.558 | 31 | 20 |
| 20.0 | 43.7 (33.7–54.1) | 94.0 (90.2–96.4) | 73.1 (59.7–83.2) | 81.7 (76.7–85.9) | 0.377 | 49 | 14 |
| **GRE ($n=420$, $PH=18$ [4.3%]) — AUC 0.943 (0.912–0.974)** |  |  |  |  |  |  |  |
| **Threshold**<br>**(mL)** | **Sensitivity**<br>**% (95% CI)** | **Specificity**<br>**% (95% CI)** | **PPV**<br>**% (95% CI)** | **NPV**<br>**% (95% CI)** | **Youden**<br>**Index** | **FN**<br>**(missed**<br>**PH)** | **FP**<br>**(ECASS**<br>**0–2)** |
| 0.1 | 94.4 (74.2–99.0) | 80.1 (75.9–83.7) | 17.5 (11.2–26.3) | 99.7 (98.3–99.9) | 0.745 | 1 | 80 |
| 0.5 | 94.4 (74.2–99.0) | 84.1 (80.2–87.3) | 21.0 (13.5–31.1) | 99.7 (98.3–99.9) | 0.785 | 1 | 64 |
| 1.0 | 94.4 (74.2–99.0) | 87.8 (84.2–90.7) | 25.8 (16.7–37.4) | 99.7 (98.4–100.0) | 0.823 | 1 | 49 |
| 1.5 | 94.4 (74.2–99.0) | 90.3 (87.0–92.8) | 30.4 (19.9–43.3) | 99.7 (98.5–100.0) | 0.847 | 1 | 39 |
| **1.6 \*** | **94.4 (74.2–99.0)** | **91.0 (87.9–93.5)** | **32.1 (21.1–45.5)** | **99.7 (98.5–100.0)** | **0.855** | **1** | **36** |
| 2.0 | 88.9 (67.2–96.9) | 92.8 (89.8–94.9) | 35.6 (23.2–50.2) | 99.5 (98.1–99.9) | 0.817 | 2 | 29 |
| 3.0 | 72.2 (49.1–87.5) | 95.3 (92.7–97.0) | 40.6 (25.5–57.7) | 98.7 (97.0–99.4) | 0.675 | 5 | 19 |

| 5.0 | 66.7 (43.7–83.7) | 97.8 (95.8–98.8) | 57.1 (36.5–75.5) | 98.5 (96.8–99.3) | 0.644 | 6 | 9 |
| --- | --- | --- | --- | --- | --- | --- | --- |
| 10.0 | 50.0 (29.0–71.0) | 98.5 (96.8–99.3) | 60.0 (35.7–80.2) | 97.8 (95.8–98.8) | 0.485 | 9 | 6 |
| 20.0 | 33.3 (16.3–56.3) | 99.3 (97.8–99.7) | 66.7 (35.4–87.9) | 97.1 (95.0–98.3) | 0.326 | 12 | 3 |
| **SWI (n=750, PH=60 [8.0%]) — AUC 0.953 (0.933–0.973)** |  |  |  |  |  |  |  |
| **Threshold**<br>**(mL)** | **Sensitivity**<br>**% (95% CI)** | **Specificity**<br>**% (95% CI)** | **PPV**<br>**% (95% CI)** | **NPV**<br>**% (95% CI)** | **Youden**<br>**Index** | **FN**<br>**(missed**<br>**PH)** | **FP**<br>**(ECASS**<br>**0–2)** |
| 0.1 | 98.3 (91.1–99.7) | 64.1 (60.4–67.6) | 19.2 (15.2–24.0) | 99.8 (98.7–100.0) | 0.624 | 1 | 248 |
| 0.5 | 98.3 (91.1–99.7) | 72.5 (69.0–75.7) | 23.7 (18.8–29.4) | 99.8 (98.9–100.0) | 0.708 | 1 | 190 |
| 1.0 | 96.7 (88.6–99.1) | 80.3 (77.2–83.1) | 29.9 (23.9–36.7) | 99.6 (98.7–99.9) | 0.770 | 2 | 136 |
| 1.5 | 96.7 (88.6–99.1) | 84.9 (82.1–87.4) | 35.8 (28.8–43.4) | 99.7 (98.8–99.9) | 0.816 | 2 | 104 |
| **1.6 \*** | **96.7 (88.6–99.1)** | **85.5 (82.7–87.9)** | **36.7 (29.6–44.5)** | **99.7 (98.8–99.9)** | **0.822** | **2** | **100** |
| 2.0 | 93.3 (84.1–97.4) | 87.4 (84.7–89.7) | 39.2 (31.5–47.3) | 99.3 (98.3–99.7) | 0.807 | 4 | 87 |
| 3.0 | 88.3 (77.8–94.2) | 90.3 (87.9–92.3) | 44.2 (35.6–53.1) | 98.9 (97.7–99.5) | 0.786 | 7 | 67 |
| 5.0 | 78.3 (66.4–86.9) | 94.3 (92.4–95.8) | 54.7 (44.2–64.7) | 98.0 (96.7–98.9) | 0.727 | 13 | 39 |
| 10.0 | 56.7 (44.1–68.4) | 97.7 (96.3–98.6) | 68.0 (54.2–79.2) | 96.3 (94.6–97.5) | 0.543 | 26 | 16 |
| 20.0 | 30.0 (19.9–42.5) | 99.1 (98.1–99.6) | 75.0 (55.1–88.0) | 94.2 (92.3–95.7) | 0.291 | 42 | 6 |

Performance metrics at fixed volume thresholds and Youden-optimal threshold (highlighted). Values are % (95% CI).
\* Optimal threshold determined by maximizing the Youden index (Sensitivity + Specificity – 1).
Abbreviations: AUC, area under the receiver operating characteristic curve; PH, parenchymal hemorrhage (ECASS $\geq 3$ ); PPV, positive predictive value; NPV, negative predictive value; FN, false negative; FP, false positive; CI, confidence interval.

### Diagnostic Discrepancies

Six false-negative PH cases were identified with a miss rate of 3.6% among the 165 cases. These occurrences comprised 4 cases on NCCT and 1 on GRE and 1 on SWI (Supplemental Table 4). Furthermore, 15 false positive results exceeded the optimal volumetric thresholds for PH among the 873 patients without hemorrhage which represented a rate of 1.7% (Supplemental Table 5). Probable causes for misclassification included large thrombi or chronic hemorrhages on GRE and SWI, and metallic artifacts or meningiomas on NCCT.

### Association Between AI-Derived Hemorrhage Volume and Functional Outcome

Three-month modified Rankin Scale (mRS) data were available for 1,488 of 1,490 patients (99.9%). AI-derived hemorrhage volume correlated with 3-month mRS (Spearman ρ = 0.353, P < 0.001; Figure 5A). A dose-response relationship was observed: good functional outcome (mRS 0–2) decreased progressively from 61.8% among patients with zero AI-detected volume (n=924) to 48.1% (0–1.6 mL, n=235), 33.0% (1.6–10 mL, n=188), 10.8% (10–50 mL, n=111), and 6.7% (>50 mL, n=30), while 3-month mortality increased correspondingly from 6.2% to 46.7% (Table 4). This gradient was consistent across all three imaging modalities (NCCT: ρ = 0.369; GRE: ρ = 0.372; SWI: ρ = 0.282; all P < 0.001). Functional outcomes stratified by expert-determined ECASS grade showed that 3-month mortality was 5.4% for ECASS 0, 14.3% for HI-1, 12.9% for HI-2, 18.3% for PH-1, and 44.6% for PH-2, with good functional outcome achieved in only 10.3% of PH cases compared with 39.5% for HI and 64.8% for no hemorrhage (Table 4).

**Figure 5.**
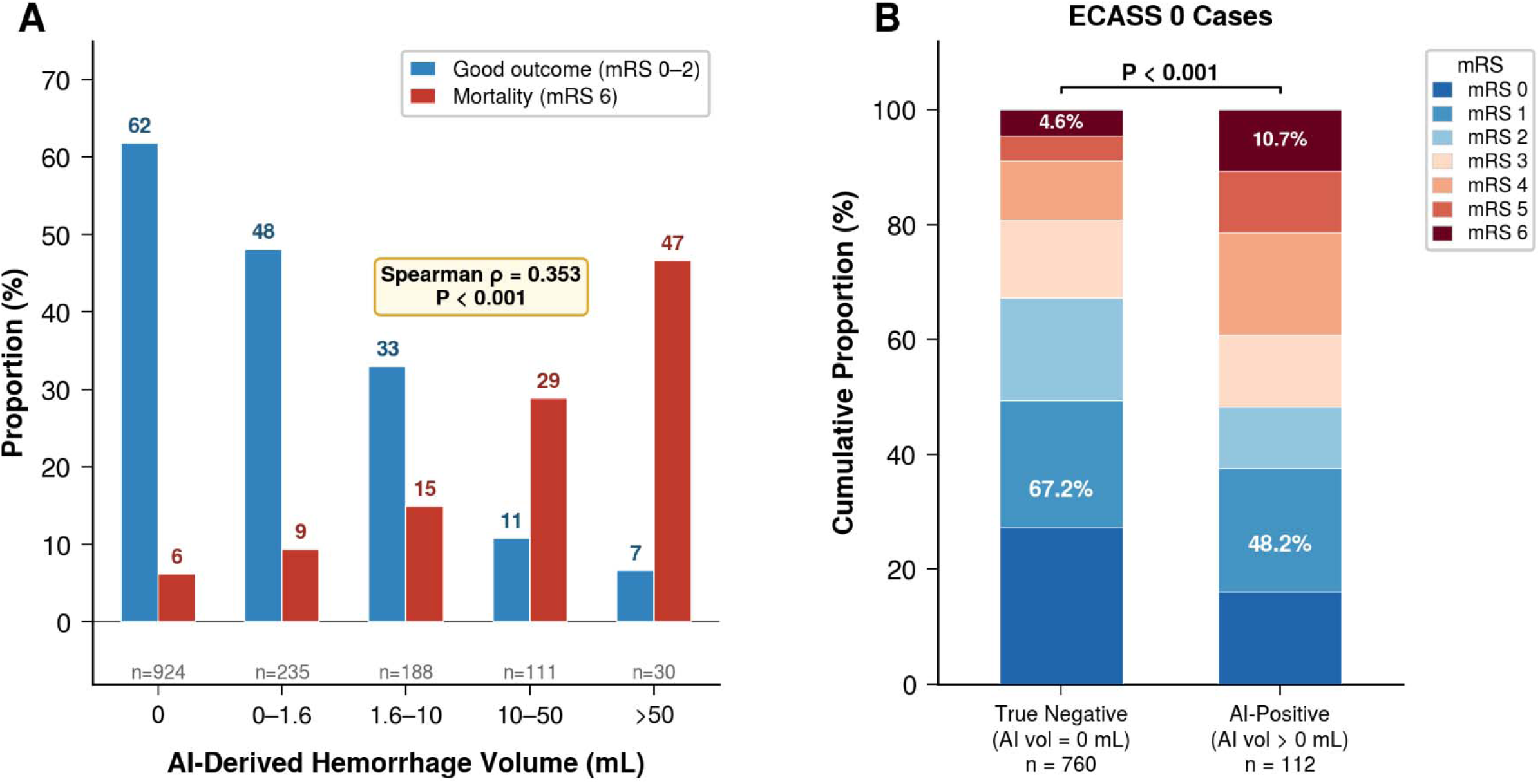
Association between AI-derived hemorrhage volume and 3-month functional outcome. A: Dose-response relationship between AI-derived hemorrhage volume categories and functional outcome. Good outcome (modified Rankin Scale [mRS] 0–2) decreases monotonically from 61.8% at 0 mL to 6.7% at >50 mL, while mortality increases from 6.2% to 46.7% (Spearman ρ = 0.353, P < 0.001). Panel B: Among patients classified as ECASS 0 (no hemorrhage) by expert consensus, AI-positive cases (volume > 0 mL) demonstrated significantly worse 3-month outcomes than true-negative cases (good outcome 48.2% vs 67.2%, mortality 10.7% vs 4.6%, P < 0.001), suggesting that automated volumetric analysis may detect prognostically relevant sub-threshold changes not captured by categorical grading. Abbreviations: AI, artificial intelligence; ECASS, European Cooperative Acute Stroke Study; mRS, modified Rankin Scale.

**Table 4.** Association Between AI-Derived Hemorrhage Volume and 3-Month Functional Outcome.

|  | N | Median mRS | Good Outcome (mRS 0–2) | Poor Outcome (mRS 3–5) | Mortality (mRS 6) |
| --- | --- | --- | --- | --- | --- |
| **By AI-Derived Hemorrhage Volume** |  |  |  |  |  |
| 0 | 924 | 2 | 571 (61.8) | 296 (32.0) | 57 (6.2) |
| 0–1.6 | 235 | 3 | 113 (48.1) | 100 (42.6) | 22 (9.4) |
| 1.6–10 | 188 | 4 | 62 (33.0) | 98 (52.1) | 28 (14.9) |
| 10–50 | 111 | 5 | 12 (10.8) | 67 (60.4) | 32 (28.8) |
| >50 | 30 | 5 | 2 (6.7) | 14 (46.7) | 14 (46.7) |
| *Spearman $\rho = 0.353$, $P < 0.001$ (overall); NCCT $\rho = 0.369$; GRE $\rho = 0.372$; SWI $\rho = 0.282$ (all $P < 0.001$)* |  |  |  |  |  |
| **By ECASS Hemorrhagic Transformation Grade** |  |  |  |  |  |
| ECASS 0 (No HT) | 872 | 2 | 565 (64.8) | 260 (29.8) | 47 (5.4) |
| ECASS 1 (HI-1) | 196 | 2 | 98 (50.0) | 70 (35.7) | 28 (14.3) |
| ECASS 2 (HI-2) | 255 | 4 | 80 (31.4) | 142 (55.7) | 33 (12.9) |
| ECASS 3 (PH-1) | 109 | 4 | 14 (12.8) | 75 (68.8) | 20 (18.3) |
| ECASS 4 (PH-2) | 56 | 5 | 3 (5.4) | 28 (50.0) | 25 (44.6) |
| **ECASS 0 Cases Stratified by AI Detection Status** |  |  |  |  |  |
| True Negative (AI vol = 0 mL) | 760 | 2 | 511 (67.2) | 214 (28.2) | 35 (4.6) |
| AI-Positive (AI vol > 0 mL) | 112 | 3 | 54 (48.2) | 46 (41.1) | 12 (10.7) |
| *Mann-Whitney $P < 0.001$* |  |  |  |  |  |

Values are n (%) unless otherwise indicated. Good outcome defined as mRS 0–2 at 3 months. Mortality defined as mRS 6. Three-month mRS data available for 1,488 of 1,490 patients (99.9%). Abbreviations: mRS, modified Rankin Scale; ECASS, European Cooperative Acute Stroke Study; HI, hemorrhagic infarction; PH, parenchymal hemorrhage.

### Clinical Significance of AI-Positive Findings Among no-HT Cases

Among 872 patients classified as ECASS 0 by expert consensus, the AI algorithm produced positive results (volume > 0 mL) in 112 cases (12.8%). These AI-positive ECASS 0 patients demonstrated significantly worse 3-month functional outcomes compared with true-negative cases (Figure 5B): good outcome (mRS 0–2) was 48.2% versus 67.2%, and mortality was 10.7% versus 4.6% (Mann-Whitney P < 0.001).

## DISCUSSION

To date, no systematic evaluation has established whether AI-based hemorrhage quantification, developed and validated for spontaneous intracranial hemorrhage in emergency settings, retains diagnostic performance in the radiographically distinct context of post-EVT hemorrhagic transformation. The present multicenter study, drawing on 1490 imaging studies across three modalities, including NCCT, GRE, and SWI, within a prospective nationwide multicenter clinical and imaging registry, addresses this gap and further demonstrates that AI-derived volumetric output carries prognostic information beyond what categorical ECASS grading captures.

The central diagnostic finding, sensitivity exceeding 94% for PH across NCCT, GRE and SWI, with AUC values of 0.900 to 0.953, warrants juxtaposition with prior evaluations of commercial AI in the post-EVT setting. [12,19] Endler and colleagues reported 95.9% sensitivity but a 15.4% false-positive rate on 495 post-EVT CT scans [12]. The NCCT algorithm evaluated here, trained on over 80,000 multi-institutional examinations [9,19], yielded comparable sensitivity (95.4%) for PH with a false-positive rate of 16.2% among those without HT on NCCT cases, consistent with the well-documented confound of iodinated contrast staining in the acute post-procedural setting. [11,20] The attenuated sensitivity for any-grade HT (61.8–79.6%) relative to PH (94.4–98.3%) reflects not algorithmic insufficiency but the ineluctable relationship between lesion volume and detectability: HI-1 petechial hemorrhages may subtend mere voxels, at or beneath the spatial resolution of segmentation algorithm of imaging modality especially for NCCT. [21] Given the difference of 3-month mortality of 5.4% for no HT, 13.5% for HI, and 27.3% for PH, HI-1 and HI-2 may be largely benign sequelae of reperfusion, whereas PH carries unequivocal prognostic weight. [3,4,5,22] That outcomes among algorithmically missed HI-1 cases did not differ from detected ones confirms negligible clinical consequence. The algorithms are thus calibrated, coincidentally, to the prognostic architecture of HT: near-complete detection of PH-2 (98.2%, mortality 44.6%) alongside permissible insensitivity to innocuous petechiae.

The superior discriminative performance of MRI-based modalities (AUC values, 0.943 from GRE and 0.953 from SWI) over NCCT (0.900) has a direct mechanistic basis. Susceptibility-sensitive sequences exploit the paramagnetism of deoxyhemoglobin and hemosiderin, rendering hemorrhagic foci conspicuous to both human and algorithmic observers [13,23], whereas NCCT attenuation differences grow equivocal amid contrast residua, edema, and partial-volume effects [11,24]. This finding reinforces the rationale for MRI-based follow-up where institutional capacity permits. [14,25] It has a practical implication for threshold application: the lower specificity of SWI at the Youden-optimal threshold (64.1%) reflects susceptibility blooming beyond the lesion boundary and argues for volumetric thresholding rather than binary detection when SWI is the follow-up modality.

The prognostically distinct behavior of AI-positive cases within the no-HT group represents the most consequential findings of this study, and it merits careful interpretation. Of 872 patients classified as ECASS 0 by expert consensus, 112 (12.8%) produced AI-positive results and exhibited substantially worse 3-month outcomes (good outcome 48.2% vs 67.2%, mortality 10.7% vs 4.6%, P < 0.001). Some of these detections reflect artifacts, including residual contrast, metallic susceptibility artifacts, or pre-existing lesions, as documented in Supplementary Table 5. Yet, the magnitude and consistency of the outcome differential across this subgroup exceed what artifact contamination alone would produce. A more parsimonious interpretation is that automated volumetry might detect sub-threshold hemorrhagic or ischemic tissue perturbations that fall beneath the perceptual resolution of categorical grading. The classical classification systems impose discrete ordinal boundaries upon a phenomenon that is biologically continuous. Under this interpretation, AI-derived volume, operating without such boundaries, may recover prognostic signal that categorical classification discards. The dichotomy of correct versus incorrect AI detection, benchmarked against expert grading as the sole reference, may thus inadequately capture the prognostic information encoded in continuous volumetric measurements.

None of the three algorithms was trained on hemorrhagic transformation *per se*. The NCCT model was developed for spontaneous ICH [9,19], and the GRE and SWI models were trained on all-cause hemorrhage spanning post-stroke, post-surgical, and traumatic etiologies. That robust performance obtains in the post-EVT domain, a context absent from any training corpus, indicates that learned hemorrhage representations generalize across pathological contexts without domain-specific retraining. The practical implication is that regulatory-cleared tools already deployed for emergency ICH triage can be evaluated for post-procedural monitoring without modification, and the metrics reported here represent a conservative estimate of what purpose-built, post-EVT-specific models might achieve.

This study has several limitations that delineate its scope. The retrospective design and non-randomized modality assignment introduce selection bias. The preponderance of SWI studies (50.3%) likely reflects institutional MRI availability, and the significant baseline differences across groups (Table 1) preclude unqualified cross-modality comparison. The absence of domain-specific training, while recast above as evidence of generalizability, simultaneously implies that reported metrics constitute a lower bound. The absence of dual-energy CT data prevents definitive hemorrhage-contrast differentiation in NCCT cases [24]. The ECASS reference standard, though applied with high inter-rater agreement, carries inherent ordinal imprecision for a continuous phenomenon. Finally, the Youden-optimal thresholds reported here were derived and evaluated within the same cohort and should not be applied to populations with ethnically and clinically different hemorrhage prevalence or imaging protocol distributions without external validation.

Automated volumetry offers a scalable, observer-independent adjunct to manual HT classification for registries and clinical trials. [21,26,27,28] The dose-response association between AI-derived volume and 3-month mRS furnishes empirical warrant for continuous volumetric endpoints, which may offer superior statistical power over ordinal categorical outcomes in future intervention trials. [29] The uniformly high PH sensitivity across modalities suggests potential utility as an autonomous triage instrument, though prospective workflow integration studies are required before clinical implementation [30]. The logical next investigative frontiers are prospective external validation of the reported thresholds, development of post-EVT–specific training data to improve sensitivity for lower-grade HT, and examination of whether serial AI-derived volumetry captures hemorrhage dynamics with prognostic implications beyond single time-point measurement.

## Statements and Declarations

## Acknowledgements

We thank the research coordinators and stroke nurses at all participating centers for their contributions to data collection.

## Author contributions (CRediT)

All authors contributed to the study concept and design, interpretation of results, and critical revision of the manuscript. W.S.R. was responsible for conceptualization, methodology, software development, formal analysis, and supervision. W.S.R. and B.J.K. drafted the original manuscript. W.S.R. and B.J.K. conducted the data visualization. B.J.K. led the supervision, funding acquisition, and project administration. H.J.H. performed data labeling and validation. M.J.L. contributed to project administration. L.S., K.K., J.G.K., S.J.L., J.K.C., T.H.P., J.Y.L., K.L., D.H.K., J.L., H.K.P., K.S.H., M.L., M.S.O., K.H.Y., D.S.G., D.E.K., H.K., J.T.K., J.G.K., J.C.C., W.J.K., J.H.K., K.S.Y., D.I.S., J.H.H., S.I.S., S.H.L., C.K., H.B.J., K.Y.P., K.J.L., C.K.K., J.K., J.Y.K., and H.J.B. contributed to data acquisition and curation. All authors had full access to the data, reviewed the final manuscript, and approved it for submission.

## Conflicts of interest

Wi-Sun Ryu, Dongmin Kim, and Myungjae Lee are employees of JLK Inc., Republic of Korea. Hee-Joon Bae reports grants from Amgen Korea Limited, Bayer Korea, Bristol Myers Squibb Korea, Celltrion, Dong-A ST, Otsuka Korea, Samjin Pharm, and Takeda Pharmaceuticals Korea Co., Ltd.; and personal fees from Amgen Korea, Bayer, Daewoong Pharmaceutical Co., Ltd., Daiichi Sankyo, Eisai Korea, Inc., JW Pharmaceutical, SK Chemicals, and Otsuka Korea, outside the submitted work. In addition, Hee-Joon Bae and Dong-Eog Kim hold stock in JLK Inc., the developer of the software evaluated in this study. Authors affiliated with JLK Inc. were excluded from data analysis and reference standard adjudication. Other authors reported no conflict of interest.

## Funding

### Data availability

Anonymized data supporting the findings of this study are available from the corresponding author upon reasonable request, subject to institutional data-sharing agreements and privacy regulations.

### Ethical approval

The study was approved by the institutional review boards of all participating centers (lead site: Seoul National University Bundang Hospital IRB [IRB No. B-2308-845-302]). Written informed consent was obtained from all participants or their legal representatives.

## Notes

### Funding Statement

This study did not receive any funding.

