## Supplementary Material for "Automated Detection and Quantification of Hemorrhagic Transformation After Endovascular Thrombectomy"

Ryu et al.

### Supplementary Methods. Development of GRE and SWI Hemorrhage Segmentation Models

#### Image Preprocessing

Brain extraction was performed using SynthStrip (Hoopes et al., *NeuroImage* 2022), followed by cropping to the non-zero bounding box. Each volume was resampled to a modality-specific target spacing derived from the median voxel dimensions of the training dataset: (4.0, 0.43, 0.43) mm for GRE and (1.0, 0.54, 0.54) mm for SWI (Supplementary Table 1). Resampling used third-order spline interpolation for images and nearest-neighbor interpolation for labels. All intracranial voxels were then z-score normalized (zero mean, unit variance). Preprocessed volumes were partitioned into 3D patches targeting approximately 512 mm isotropic spatial coverage, yielding patch sizes of (28, 224, 256) voxels for GRE and (80, 160, 192) voxels for SWI, extracted with 50% overlap in all dimensions.

#### Network Architecture

Both models employed the nnU-Net 3D full-resolution configuration (Isensee et al., *Nat Methods* 2021), a fully convolutional encoder–decoder with skip connections. The encoder comprises two (1,3,3) convolution layers followed by five (3,3,3) convolution stages with strided downsampling; each layer is followed by instance normalization and Leaky ReLU. The decoder applies transposed convolutions for upsampling, concatenates encoder features via skip connections, and passes them through localization blocks of five (3,3,3) and one (1,3,3) convolution layers (inverse encoder order). The output layer produces four-channel voxel-wise predictions (background, hemorrhage, siderosis, cerebral microbleeds) via softmax activation.

#### Training Data and Annotation

The GRE model was trained on 1,441 studies from 12 institutions (hemorrhage-positive: 34.3%) and the SWI model on 1,152 studies from 12 institutions (hemorrhage-positive: 29.1%), spanning three MRI manufacturers and both 1.5 T and 3.0 T field strengths (Supplementary Table 2). Voxel-level annotations were generated under a multi-label schema (hemorrhage, superficial siderosis, cerebral microbleeds) and independently reviewed and adjudicated by two board-certified vascular neurologists with expertise in stroke neuroimaging.

**Training Strategy**

Models were trained with a combined Dice and binary cross-entropy loss, optimized by Adam (learning rate $3\times10^{-4}$, weight decay $3\times10^{-5}$) with ReduceLROnPlateau scheduling and batch size 2. Each epoch comprised 250 randomly sampled mini-batches rather than a full dataset pass. Positive patches were oversampled at a 1:2 positive-to-negative ratio. Online augmentation included elastic deformation, scaling, rotation, mirroring, and gamma correction. Early stopping was triggered after 50 iterations without validation loss improvement; the GRE and SWI models converged at epochs 433 and 442, respectively. The entire training set served as the validation set for early stopping; the combination of stochastic patch cropping, augmentation, and mini-batch sub-sampling provided sufficient regularization, as confirmed by concordant external validation performance (NVIDIA A6000; Python 3.10.4; PyTorch 2.0.1; CUDA 12.0).

**CSF-Guided Post-Processing**

A separately trained nnU-Net 2D model generated CSF masks for each axial slice (Supplementary Table 1). For each connected component labeled as hemorrhage or siderosis by the primary model, the proportion of voxels overlapping with CSF space was computed; components with ≥50% CSF overlap were reclassified as superficial siderosis and excluded from the hemorrhage volume.

**Inference and Volume Calculation**

All studies were processed in fully automated batch mode. Overlapping 3D patches were extracted, passed through the segmentation model, and augmented at test time via axial mirroring. Patch predictions were aggregated using Gaussian-weighted averaging. After CSF-guided reclassification, total hemorrhage volume (mL) was computed as the product of hemorrhage-labeled voxel count and physical voxel volume.

**External Validation**

The GRE model was externally validated on 310 studies from 2 institutions not represented in training (hemorrhage-positive: 39.7%); the SWI model on 216 studies from 2 independent institutions (hemorrhage-positive: 15.3%). Annotations followed the same dual-expert protocol. Case-averaged voxel-level metrics were: GRE DSC 0.938, sensitivity 0.951, specificity 0.957; SWI DSC 0.862, sensitivity 0.970, specificity 0.885 (Supplementary Table 1).

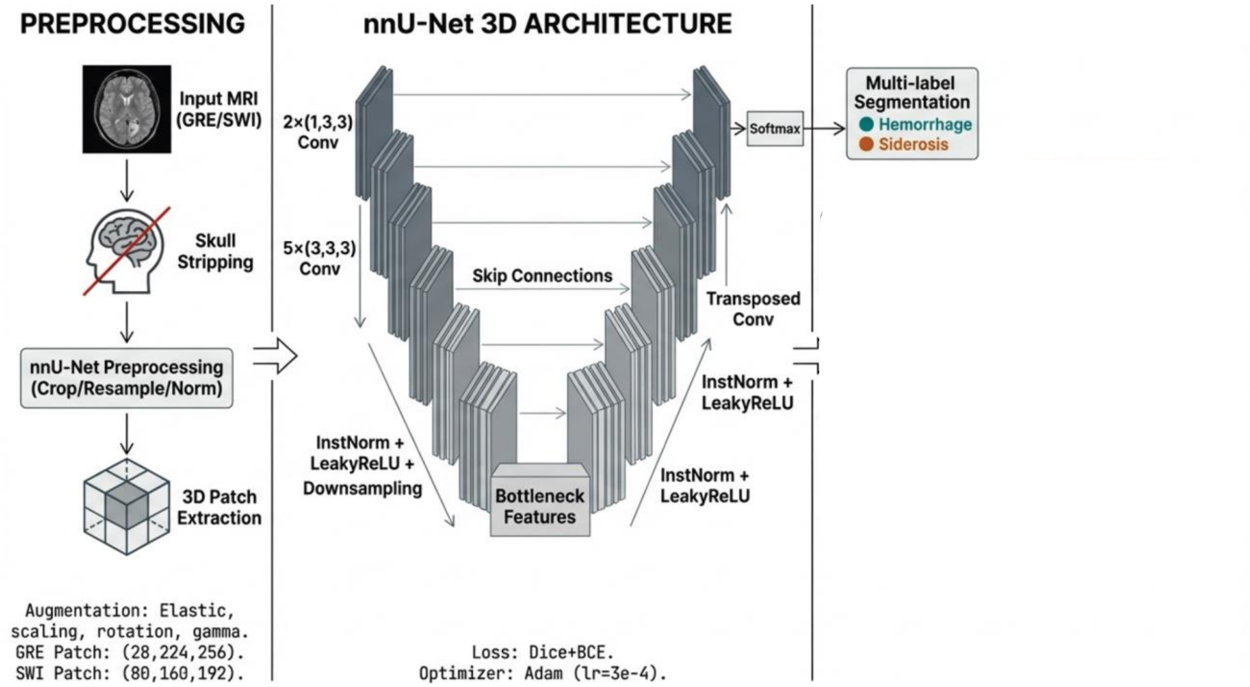


**Supplementary Figure 1. AI hemorrhage segmentation pipeline for GRE and SWI sequences.**

The pipeline consists of three stages: (1) preprocessing including SynthStrip skull stripping, nnU-Net resampling and Z-score normalization, and 3D patch extraction with 50% overlap; (2) a fully convolutional encoder–decoder network (nnU-Net 3D) with skip connections, trained using Dice + binary cross-entropy loss, producing multi-label segmentation of hemorrhage, superficial siderosis, and cerebral microbleeds; and (3) post-processing using a separately trained CSF segmentation model (nnU-Net 2D) to classify lesions as siderosis (≥50% spatial overlap with CSF) or parenchymal hemorrhage. The final output consists of a binary hemorrhage detection result and continuous volumetric measurement in milliliters. Model-specific parameters are shown: GRE (target spacing 4.0 × 0.43 × 0.43 mm, patch size 28 × 224 × 256 voxels) and SWI (target spacing 1.0 × 0.54 × 0.54 mm, patch size 80 × 160 × 192 voxels). Abbreviations: CSF, cerebrospinal fluid; GRE, gradient-recalled echo; SWI, susceptibility-weighted imaging.

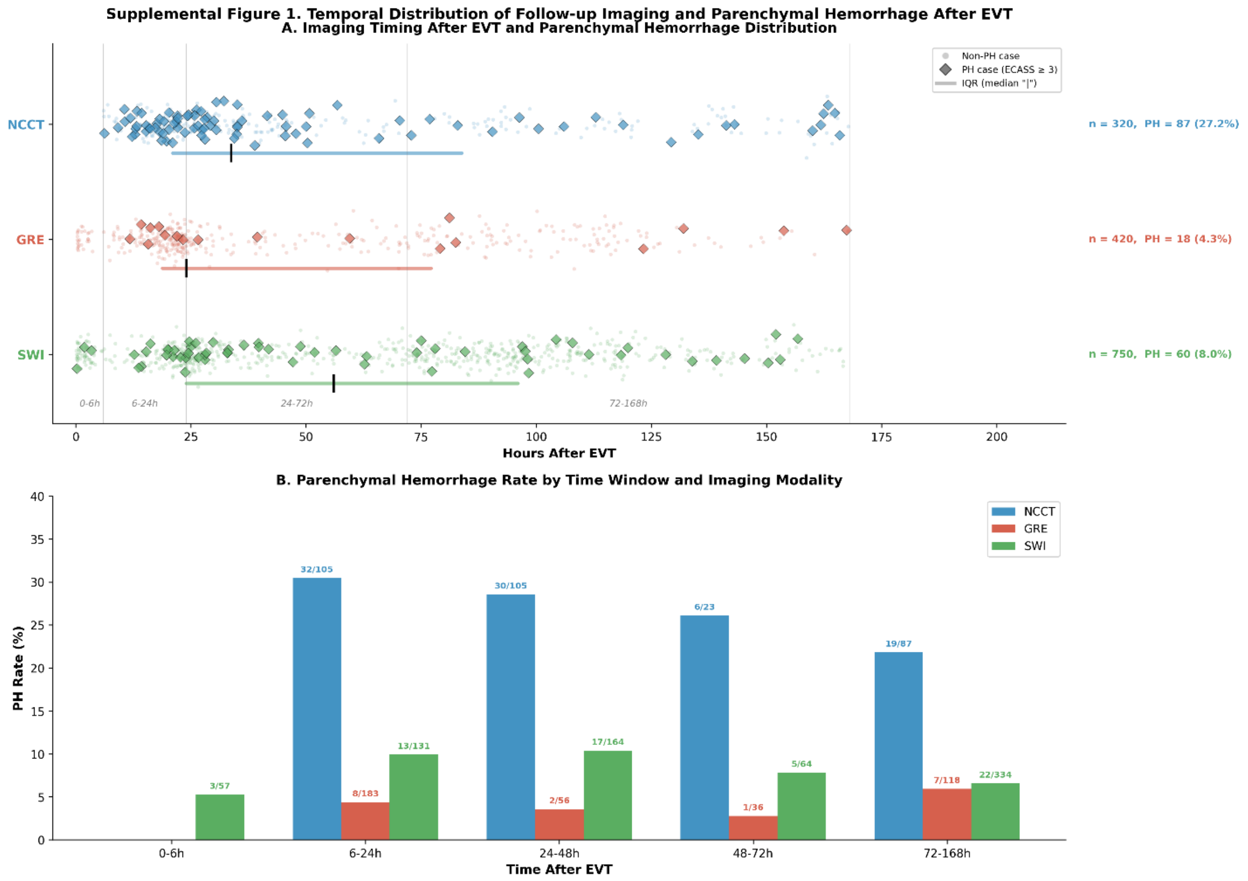


**Supplementary Figure 2. Temporal distribution of follow up neuroimaging after endovascular thrombectomy stratified by the presence of parenchymal hemorrhage.**

The box plots illustrate the interval in hours from the index procedure to the acquisition of follow up imaging for each modality. The cohort is partitioned into patients with parenchymal hemorrhage and those without such findings. Horizontal lines within the boxes represent median values and the boundaries signify the interquartile range. NCCT was obtained at a median interval of 33.7 hours while SWI was utilized at a median of 56.0 hours. These data demonstrate the temporal prioritization of computed tomography during the hyperacute post procedural period. Abbreviations: EVT, endovascular thrombectomy; GRE, gradient-recalled echo; NCCT, non-contrast computed tomography; PH, parenchymal hemorrhage; SWI, susceptibility-weighted imaging.

**Supplemental Table 1. Development and Validation of GRE and SWI Hemorrhage Segmentation Models**

| | GRE Model | SWI Model |
|---|---|---|
| **Architecture & Framework** | | |
| Base framework | nnU-Net 3D | nnU-Net 3D |
| Network type | Encoder–decoder with skip connections | Encoder–decoder with skip connections |
| Segmentation labels | Hemorrhage, Siderosis, CMB (multi-label) | Hemorrhage, Siderosis, CMB (multi-label) |
| Skull stripping | SynthStrip | SynthStrip |
| **Training Data** | | |
| No. of studies | 1,441 | 1,152 |
| No. of institutions | 12 | 12 |
| Hemorrhage-positive, n (%) | 494 (34.3%) | 335 (29.1%) |
| **External Validation** | | |
| No. of studies | 310 | 216 |
| No. of institutions | 2 | 2 |
| Hemorrhage-positive, n (%) | 123 (39.7%) | 33 (15.3%) |
| **Preprocessing** | | |
| Target spacing (mm) | (4.0, 0.43, 0.43) | (1.0, 0.54, 0.54) |
| Patch size (voxels) | (28, 224, 256) | (80, 160, 192) |
| Intensity normalization | Z-score | Z-score |
| **Training Hyperparameters** | | |
| Loss function | Dice + BCE | Dice + BCE |
| Optimizer | Adam (lr = $3\times10^{-4}$, wd = $3\times10^{-5}$) | Adam (lr = $3\times10^{-4}$, wd = $3\times10^{-5}$) |
| Scheduler | ReduceLROnPlateau | ReduceLROnPlateau |
| Batch size | 2 | 2 |
| Early stopping | 50 iterations w/o improvement | 50 iterations w/o improvement |
| Final training epoch | 433 | 442 |
| Data augmentation | Elastic deform., scaling, rotation, mirroring, gamma | Elastic deform., scaling, rotation, mirroring, gamma |
| **CSF Classification** | | |
| CSF model framework | nnU-Net 2D | nnU-Net 2D |
| CSF target spacing (mm) | (4.0, 0.45, 0.45) | (1.0, 0.31, 0.31) |
| CSF patch size (pixels) | (320, 384) | (448, 512) |
| Classification rule | ≥50% CSF overlap → Siderosis | ≥50% CSF overlap → Siderosis |
| **Segmentation Performance** | | |
| Dice similarity coefficient | 0.938 | 0.862 |
| Sensitivity (voxel-level) | 0.951 | 0.970 |
| Specificity (voxel-level) | 0.957 | 0.885 |
| **Computing Environment** | | |
| Software | Python 3.10.4, PyTorch 2.0.1 | Python 3.10.4, PyTorch 2.0.1 |
| Hardware | NVIDIA A6000, CUDA 12.0 | NVIDIA A6000, CUDA 12.0 |

*Abbreviations: BCE, binary cross-entropy; CMB, cerebral microbleed; CSF, cerebrospinal fluid; DSC, Dice similarity coefficient.*

### Supplemental Table 2. Imaging Parameter Distribution of Training and External Validation Datasets

| | SWI Training (n=1,152) | SWI External (n=216) | GRE Training (n=1,441) | GRE External (n=310) |
|---|---|---|---|---|
| **MRI Manufacturer** | | | | |
| Philips | 609 | 66 | 763 | 0 |
| Siemens | 52 | 150 | 277 | 0 |
| GE | 41 | 0 | 364 | 0 |
| Unknown | 0 | 0 | 37 | 310 |
| **Magnetic Field Strength** | | | | |
| 1.5 T | 5 | 23 | 705 | 307 |
| 3.0 T | 1,147 | 193 | 735 | 3 |
| **Pixel Spacing (mm)** | | | | |
| < 0.3 | 412 | 62 | 6 | 0 |
| 0.3–0.5 | 535 | 129 | 1,421 | 307 |
| 0.5–0.7 | 115 | 24 | 6 | 4 |
| ≥ 0.7 | 90 | 1 | 8 | 0 |
| **Slice Spacing (mm)** | | | | |
| < 2 | 830 | 135 | 0 | 0 |
| 2–3 | 252 | 81 | 0 | 0 |
| 3–5 | 58 | 0 | 312 | 76 |
| ≥ 5 | 12 | 0 | 1,129 | 234 |
| **Slice Thickness (mm)** | | | | |
| < 2 | 91 | 26 | 0 | 0 |
| 2–3 | 762 | 190 | 0 | 0 |
| 3–5 | 287 | 0 | 312 | 76 |
| ≥ 5 | 12 | 0 | 1,129 | 234 |

*Values represent number of studies. Pixel spacing and slice parameters were binned from continuous values for presentation. Abbreviations: GRE, gradient-recalled echo; SWI, susceptibility-weighted imaging.*

**Supplemental Table 3. Baseline Characteristics by ECASS Hemorrhagic Transformation Grade**

| | **ECASS 0 (n=873)** | **ECASS 1 (n=196)** | **ECASS 2 (n=256)** | **ECASS 3 (n=109)** | **ECASS 4 (n=56)** | **P value** |
|---|---|---|---|---|---|---|
| Age, years | 70 (61–80) | 75.5 (63.8–82) | 74.5 (64–81.2) | 75 (66–82) | 78.5 (64–84) | <0.001 |
| Male sex, n (%) | 523 (59.9) | 114 (58.2) | 119 (46.5) | 64 (58.7) | 35 (62.5) | 0.039 |
| NIHSS at admission | 7 (4–12) | 12 (6–16.2) | 13 (9–18) | 15 (11–18) | 15 (10–19) | <0.001 |
| Pre-stroke mRS | 0 (0–0) | 0 (0–0) | 0 (0–1) | 0 (0–0) | 0 (0–0) | 0.003 |
| Comorbidities | | | | | | |
| Hypertension, n (%) | 523 (59.9) | 131 (66.8) | 165 (64.5) | 60 (55.0) | 38 (67.9) | 0.459 |
| Diabetes mellitus, n (%) | 226 (25.9) | 50 (25.5) | 95 (37.1) | 30 (27.5) | 19 (33.9) | 0.067 |
| Atrial fibrillation, n (%) | 252 (28.9) | 83 (42.3) | 144 (56.2) | 62 (56.9) | 32 (57.1) | <0.001 |
| Dyslipidemia, n (%) | 322 (36.9) | 67 (34.2) | 92 (35.9) | 31 (28.4) | 22 (39.3) | 0.831 |
| Prior stroke, n (%) | 121 (13.9) | 26 (13.3) | 55 (21.5) | 26 (23.9) | 14 (25.0) | 0.018 |
| IV thrombolysis, n (%) | 646 (74.0) | 112 (57.1) | 136 (53.1) | 60 (55.0) | 31 (55.4) | <0.001 |
| Imaging modality | | | | | | |
| NCCT, n (%) | 117 (13.4) | 28 (14.3) | 88 (34.4) | 50 (45.9) | 37 (66.1) | |
| GRE, n (%) | 310 (35.5) | 51 (26.0) | 41 (16.0) | 12 (11.0) | 6 (10.7) | |
| SWI, n (%) | 446 (51.1) | 117 (59.7) | 127 (49.6) | 47 (43.1) | 13 (23.2) | |
| Imaging timing, hours | 32.8 (20.5–88.2) | 62.0 (23.9–105.6) | 42.7 (24.2–96.3) | 36.1 (24.2–90.5) | 22.1 (15.6–46.8) | <0.001 |
| AI volume, mL | 0 (0–0) | 0 (0–0.9) | 1.7 (0.2–4.8) | 9.8 (3.7–18.0) | 34.2 (18.0–66.0) | <0.001 |
| 3-month mRS | 2 (0–3) | 2.5 (1–4) | 4 (2–5) | 4 (3–5) | 5 (4–6) | <0.001 |
| Good outcome (mRS 0–2), n (%) | 565 (64.7) | 98 (50.0) | 80 (31.2) | 14 (12.8) | 3 (5.4) | |

Values are presented as median (interquartile range) or number (percentage). Abbreviations: ECASS, European Cooperative Acute Stroke Study; NIHSS, National Institutes of Health Stroke Scale; mRS, modified Rankin Scale; IV, intravenous; NCCT, non-contrast computed tomography; GRE, gradient-recalled echo; SWI, susceptibility-weighted imaging; AI, artificial intelligence.

**Supplemental Table 4. Parenchymal Hemorrhage Cases Not Detected by AI (False Negatives)**

| # | Imaging Modality | ECASS Grade | Hours After EVT | Remarks |
|---|---|---|---|---|
| 1 | GRE | 4 | 79.1 | FN: Distant location hemorrhage |
| 2 | NCCT | 3 | 129.3 | FN:Iso-attenuated hemorrhage |
| 3 | NCCT | 3 | 141.2 | FN:Iso-attenuated hemorrhage |
| 4 | NCCT | 3 | 27.9 | FN:Iso-attenuated hemorrhage |
| 5 | NCCT | 3 | 164.8 | FN:Iso-attenuated hemorrhage |
| 6 | SWI | 3 | 13.6 | FN: diffuse SAH |

Cases with manual ECASS grade $\geq 3$ (PH-1 or PH-2) but AI-measured hemorrhage volume = 0 mL. Summary: 6 false-negative PH cases out of 165 total PH cases (3.6% miss rate). NCCT: 4, GRE: 1, SWI: 1. Abbreviations: AI, artificial intelligence; ECASS, European Cooperative Acute Stroke Study; EVT, endovascular thrombectomy; FN, false negative; GRE, gradient-recalled echo; NCCT, non-contrast computed tomography; PH, parenchymal hemorrhage; SAH, subarachnoid hemorrhage; SWI, susceptibility-weighted imaging.

**Supplemental Table 5. False-Positive Cases Exceeding Optimal PH Volume Threshold (ECASS 0)**

| # | Imaging Modality | Threshold (mL) | AI Volume (mL) | Hours After EVT | Remarks |
|---|---|---|---|---|---|
| 1 | GRE | 1.6 | 4.2 | 89.3 | FP:Large clot |
| 2 | GRE | 1.6 | 2.5 | 89.1 | FP:Large clot |
| 3 | GRE | 1.6 | 1.9 | 18.8 | FP: old hemorrhage |
| 4 | GRE | 1.6 | 1.7 | 14.4 | FP: old hemorrhage |
| 5 | GRE | 1.6 | 1.7 | 22.2 | FP: old hemorrhage |
| 6 | NCCT | 1.8 | 79 | 36.0 | FP: severe infarct |
| 7 | NCCT | 1.8 | 19.9 | 34.3 | FP: metal artifact |
| 8 | NCCT | 1.8 | 4.1 | 16.4 | FP: falx thickening |
| 9 | NCCT | 1.8 | 2.1 | 163.2 | FP: meningioma |
| 10 | SWI | 1.6 | 6.4 | 33.4 | FP: s/p craniotomy |
| 11 | SWI | 1.6 | 3.6 | 79.4 | FP: old hemorrhage |
| 12 | SWI | 1.6 | 3 | 109.0 | FP: old hemorrhage |
| 13 | SWI | 1.6 | 2.4 | 85.3 | FP: old hemorrhage |
| 14 | SWI | 1.6 | 1.9 | 124.7 | FP: old hemorrhage |
| 15 | SWI | 1.6 | 1.7 | 21.9 | FP: old hemorrhage |

Cases classified as ECASS 0 where AI-derived volume exceeded the modality-specific optimal threshold. Summary: 15 false-positive PH cases (1.7% of ECASS 0 cases) were identified where AI-derived volumes exceeded optimal thresholds. Major causes included large thrombi or chronic hemorrhages on MRI (n=11) and metallic artifacts or meningiomas on NCCT (n=4). Abbreviations: AI, artificial intelligence; ECASS, European Cooperative Acute Stroke Study; EVT, endovascular thrombectomy; FP, false positive; GRE, gradient-recalled echo; NCCT, non-contrast computed tomography; PH, parenchymal hemorrhage; SWI, susceptibility-weighted imaging.
